# Leveraging Large Language Models for Temporal PHI De-identification in Real-World Clinical Notes

**DOI:** 10.64898/2026.09.22.26360316

**Authors:** Xiaomeng Wang, Liwei Wang, Andrew Wen, Rui Li, Shuyu Lu, Hu Yian, Li Xin, Heather Lyu, Hongfang Liu

**Affiliations:** McWilliams School of Biomedical Informatics, University of Texas Health Science Center at Houston, Houston, TX; Department of Surgical Oncology, Division of Surgery, The University of Texas MD Anderson Cancer Center, Houston, TX

## Abstract

De-identifying temporal protected health information (PHI) is challenging because temporal expressions appear in diverse formats and must be transformed while preserving clinically meaningful timelines. This study proposes and evaluates an LLM-based framework for temporal PHI de-identification in real-world clinical notes. Using 1,148 notes from 30 sarcoma patients (28,431 annotated temporal entities), we evaluated multiple modern LLMs for temporal entity extraction and surrogate generation. Proprietary models achieved strong extraction performance, with GPT-4o achieving the best overall results. Most models preserved temporal formatting (>99%), but surrogate generation remained challenging. GPT-5.4 achieved the highest shift correctness (90.2% on a shared-entity subset) and the lowest order violations. Error analysis revealed systematic shift deviations (±1, ±30/31, ±365 days), highlighting persistent limitations in LLM temporal reasoning. These findings suggest that reliable temporal de-identification will require hybrid or multi-agent approaches beyond standalone LLMs.

## Introduction

Temporal information plays an essential role in Electronic Health Records (EHR). Clinical notes encode diagnoses, procedures, medication schedules, symptom trajectories, and follow-up events using dates, times, durations, and relative temporal expressions. Accurate interpretation of temporal information is indispensable for understanding disease progression ^1^, treatment responses ^2^, and care delivery pathways ^3^. Studies have shown that extracting temporal information from EHRs is widely used in emerging data-driven healthcare applications, such as predictive modeling ^4-6^ and digital twin systems that simulate patient clinical trajectories ^7,8^.

However, temporal information is also considered a major category of protected health information (PHI). In the United States, guidance from the U.S. Department of Health and Human Services emphasizes that dates and other time-related information associated with an individual must be removed or transformed before clinical notes can be shared for research purposes, in accordance with the Health Insurance Portability and Accountability Act (HIPAA) Privacy Rule ^9^.

Deidentifying temporal PHI is challenging because temporal information is often embedded in clinical notes and appears in diverse formats ^10,11^, including explicit dates (e.g., “03/15/2020”), relative expressions (e.g., “two weeks ago”), and partially specified references (e.g., “in Feb”). Moreover, effective temporal de-identification requires not only detecting these entities but also transforming them in a way that preserves clinically meaningful temporal relationships within a patient’s record ^12^. As clinical notes document past medical events, temporal information is not limited to the encounter date, adding complexity to temporal PHI de-identification.

Traditional approaches to clinical de-identification have relied on rule-based systems, machine learning models, or hybrid pipelines ^13-15^ trained on annotated datasets such as the i2b2 de-identification challenges ^16-20^. While these systems have achieved strong performance in identifying common PHI categories, deidentification of temporal PHI remains challenging due to its linguistic variability and ambiguity. In recent years, large language models (LLMs) have advanced rapidly in natural language processing capabilities and have demonstrated strong performance on information extraction tasks. Emerging studies suggest that LLMs can perform clinical entity recognition and PHI detection with competitive performance under zero-shot or few-shot prompting settings ^21-24^.

Due to the limited access to real-world data, the performance of LLMs for temporal PHI identification has not been widely evaluated in real clinical notes. Most prior studies have focused on benchmark datasets or general PHI extraction tasks rather than specifically examining temporal information ^16-20^. However, temporal de-identification in clinical practice extends beyond entity detection. In practice, extracted temporal entities are typically replaced with surrogate values to preserve clinically meaningful time intervals. For example, the MIMIC-III database applies a patient-level date-shifting approach, in which all dates associated with an individual patient are shifted by a consistent random offset ^25^. Despite these practices, there is limited evidence on whether LLMs can reliably generate high-quality temporal surrogates that maintain consistent shift values and preserve chronological relationships. Systematic evaluation metrics for assessing the quality of temporal surrogate generation remain underexplored. Generating temporal surrogates requires accurate date arithmetic, consistent application of transformation rules, and preservation of temporal order across multiple clinical events. Furthermore, existing studies rarely examine LLM empowered temporal de-identification as a multi-stage pipeline, where errors may arise at different stages.

To address this gap, our work focuses on the challenge of generating temporal surrogates that preserve patient-level temporal structure. Specifically, we evaluate whether LLM-generated replacements maintain formatting consistency, apply the intended temporal shift, and preserve chronological ordering across clinical notes. This study contributes a systematic evaluation framework for identifying where current LLM-based temporal de-identification succeeds and fails.

## Methods

### Data and Annotation

Our data were collected from the UTHealth Houston OMOP Common Data Model (CDM), which provides an OMOP view integrating longitudinal electronic health record (EHR) data from UTHealth’s McGovern Medical School outpatient practice, UTPhysicians (UTP). This study was approved by the Institutional Review Board of UTHealth (HSC-SBMI-13-0549).

Using this OMOP environment, we formed a sarcoma cohort ^26^ by first identifying individuals with ICD-10 diagnosis codes determined by clinical experts for sarcoma. We then confirmed sarcoma cases if the keyword “sarcoma” appeared in the patient’s clinical notes. After that, we randomly selected 30 patients from this pool. For each selected patient, we extracted all of their available clinical notes through September 2024. In total, 1,212 documents were retrieved and preprocessed for pre-annotation of deidentification using the Open Health Natural Language Processing Toolkit (OHNLPTK) with a RoBERTA model finetuned for de-identification on the i2b2 dataset [39]. The specific entities tagged in accordance with HIPAA Safe Harbor guidelines included person names, locations, organization names, age, phone numbers, email addresses, datetime, zip codes, professions, user IDs, and record identifiers (e.g., MRNs and SSNs). Human annotations were conducted on top of the pre-annotations. Specifically, two human annotators further annotated PHI information for the same 250 notes, and disagreements were resolved through discussion to reach consensus. The overall inter-annotator agreement (IAA) was 0.8564 (F1), and the IAA for temporal PHI was 0.8880, indicating a relatively high level of agreement between annotators. To reduce efforts and time, human annotators divided the remaining notes equally and completed the annotation.

As this study focuses on temporal PHI, we excluded 64 notes in which no temporal entity was identified during annotation. The final dataset, therefore, consisted of 1,148 clinical notes from 30 patients, 28,431 temporal entities. Clinical notes varied in length, with an average of 857 words per note (SD = 622). Each note contained a mean of 24.8 temporal entities (SD = 26.0, median = 15). All notes in the final dataset contained at least one temporal entity, and 76.2% of notes contained five or more temporal entities, indicating a high density of temporal information in sarcoma patient documentation. Temporal expressions appeared in a variety of formats. The most common format was numeric calendar dates (e.g., “01/01/2026; 47.96%), followed by text-based date expressions (e.g., “January 1st, 2026”; 25.15%) and date-time combinations (e.g., “01/01/2026 14:00”; 22.31%). Less frequent formats included irregular date-time expressions (e.g., “post-op day 1”; 2.88%), year-only references (e.g., “2026”; 0.81%), weekday-only references (e.g., “Saturday”; 0.75%), and time-only expressions (e.g., “4pm”; 0.14%).

### Temporal PHI De-identification Framework

#### Step 1 Temporal PHI Extraction with LLMs

We used the six models to represent complementary categories of LLMs: proprietary models with strong instruction-following and reasoning capabilities (GPT-4.1, GPT-4o, GPT-5.2, and GPT-5.4), a widely used open-weight model (LLaMA-3.3-70B), and a biomedical open-weight model (MedGemma-text). Each model was prompted with the original clinical notes and instructed to extract temporal PHI into a structured JSON format. The few-shot prompts included exactly 14 annotated examples, systematically selected to provide two examples for each of the seven temporal expression formats. All models were evaluated without additional fine-tuning.

#### Step 2 Patient-Level Temporal Shift Configuration

After extracting temporal PHI, we evaluated whether LLMs could perform surrogate generation to replace the original temporal entities for deidentification at the patient level. Since LLMs are not designed to reliably generate random numbers ^27^, we used a reproducible random seed (derived via SHA-256 hashing of the patient ID and a namespace string) to deterministically generate five candidate day-level shifts within a [-365, 365] days range, and five candidate minute-level shifts within a [-720, 720] minutes range. Specifically, the prompt provided to the models included: (a) the candidate shifts lists, (b) patient-level summary statistics (e.g., entity counts, minimum/maximum years observed), and (c) the raw extracted entities to supply clinical timeline context. Based on this information, the model was instructed to select exactly one day-level shift and one minute-level shift from the candidate lists. The prompt emphasized maintaining the temporal structure while increasing privacy protection.

#### Step 3 Entity-Level Temporal Surrogate Generation

Finally, we adopted LLMs to apply the selected patient-level shifts (Step 2) to the extracted temporal entities (Step 1) to generate temporal surrogates. For each note, we provided the LLMs with (a) information: the patient ID, the assigned day and minute shifts, and the original temporal entities; (b) instruction: shift calendar dates by the day-level offset, clock times by the minute-level offset, and combined date-time expressions by both. Throughout this process, the model was required to preserve the original surface formatting of the temporal expression as much as possible, including separator styles (e.g., slash or dash), month representations (numeric or textual), ordering conventions, zero-padding, AM/PM notation, punctuation, and surrounding spacing. This ensured that the generated surrogates remained stylistically consistent with the original clinical text while altering their temporal meaning.

### Evaluation of De-identification Performance

#### Evaluation of Temporal PHI extraction

Extraction performance was first evaluated against the gold standard annotations using standard named-entity recognition metrics: precision, recall, and F1-score. Then, to better reflect practical clinical text-extraction scenarios, we conducted a fuzzy span-matching evaluation, capturing cases where the model correctly identifies them but slightly mismatches on span boundaries. Under this setting, a predicted span was considered correct if its Jaccard similarity with the gold span exceeded 0.85. Furthermore, to analyze model behavior across different temporal structures, we conducted an error analysis stratified by entity type. Using the temporal expression formats previously defined, we computed the distribution of false negatives (FN) and false positives (FP) across models to identify challenges associated with specific temporal formats.

#### Evaluation of Temporal Surrogate Generation

The evaluation was first conducted on the full dataset (full). However, because surrogate generation depends on entities detected during the upstream PHI extraction stage, differences in detection recall across models could introduce bias. To control this, we conducted an additional evaluation using a subset of 7,811 temporal entities that were successfully recognized by all evaluated models (sample). By restricting the analysis to these shared entities, we isolated the surrogate-generation capability from entity-detection performance, enabling a fair cross-model comparison.

The correctness and reliability of temporal surrogates were examined in three dimensions: formatting consistency, ordering integrity, and temporal shifting accuracy.

##### Formatting consistency

We measured whether the structure of a temporal entity remains consistent after shifting. For each entity, we compared the structural format of the original expression with the LLM-generated surrogate using a rule-based format signature that captures separators, token types, and time indicators. A surrogate was considered format-preserving if the shifted entity’s structural format matched that of the original text.

##### Ordering integrity

We evaluated whether the chronological order of events was preserved after surrogation. For each patient, temporal entities were extracted and arranged by their appearance order in the clinical notes. Temporal order was evaluated by comparing adjacent entity pairs (*t*_*e*1_, *t*_*e*2_). The temporal relation *r*(*t*_*e*1_, *t*_*e*2_) was determined using a tolerance-based comparison:

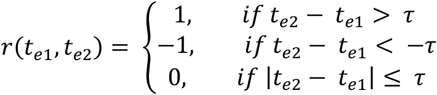

]where τ is a 1,440-minute (±1 day) tolerance window to account for minor variations. A relation value of 1 indicates that the second timestamp occurs later than the first, -1 indicates that it occurs earlier, and 0 indicates that the two timestamps fall within the tolerance window and are treated as temporally equivalent. An order violation occurs when this pairwise relation changes after surrogate generation. For example, if “03/14/2020” precedes “03/15/2020” in the original note but the outputs reverse this relation, this pair is counted as an order violation. The overall violation ratio was calculated as the proportion of adjacent pairs where the original and shifted temporal relations differed:

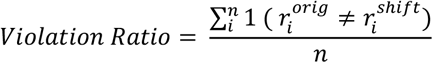

where *n* denotes the number of comparable adjacent temporal entity pairs within a patient, 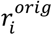 and 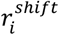 represent the temporal relations computed from the original and shifted timestamps for pair *i*, respectively, and 1(·)is an indicator function that equals 1 when the condition is true and 0 otherwise.

##### Temporal shifting accuracy

We evaluated whether the generated surrogate accurately reflected the assigned patient-level shift. Both original and shifted expressions were parsed into normalized timestamps to calculate the observed temporal offset. A shift was considered accurate if this observed offset exactly matched the expected shift. For example, suppose that the original entity is “03/15/2020” and the assigned patient-level day shift is +30 days. A correct surrogate would be “04/14/2020”. If the model instead generates “04/17/2020,” the surrogate is considered inaccurate. For entities failing this check, we computed the residual difference between the observed and expected shifts to analyze systematic model errors.

### Experimental Setup

We conducted a comparative evaluation of both proprietary and open-weight models to assess the LLM’s ability to extract temporal PHI from clinical notes. For GPT models, inference was performed through the institutionally approved, HIPAA-compliant environment via the Azure OpenAI API (version: 2025-01-01-preview) from a Python environment (Python 3.10.1). Decoding parameters (temperature = 0, top-p = 1) were fixed to ensure consistent outputs across runs. For open-weight models, inference was conducted locally using Hugging Face llama 3.3-70B and MedGemma-text-27B on an 8× NVIDIA H100 GPU server, with CUDA Version 12.7. Decoding parameters were fixed to match the GPT configuration as closely as possible. The same preprocessing, prompt format, and post-processing pipeline were applied across all models to ensure a fair comparison.

## Results

### LLM-Based Temporal PHI Extraction Evaluation

Among all evaluated models, the proprietary models consistently outperformed open-weight models in temporal PHI extraction (Table 1). GPT-4o with few-shot prompting achieved the best overall performance, while its zero-shot counterpart showed very similar results. Few-shot GPT-4.1 achieved the highest recall but showed a slightly lower overall F1 score due to reduced precision. LLaMA-3.3-70B achieved moderate results under both prompting settings, whereas MedGemma-text struggled with low recall, resulting in the lowest overall F1 scores despite improvements from few-shot prompting.

**Table 1.** Performance of LLMs for Temporal PHI Extraction under Exact and Fuzzy Span Matching

| Model | Prompt | Exact Match (Micro/Macro) |  |  | Fuzzy Match (Micro/Macro) |  |  |
| --- | --- | --- | --- | --- | --- | --- | --- |
|  |  | P | R | F1 | P | R | F1 |
| GPT-4.1 | zero-shot | 0.800 / 0.802 | 0.866 / 0.891 | 0.831 / 0.831 | 0.863 / 0.857 | 0.934 / 0.953 | 0.897 / 0.889 |
|  | few-shot | 0.841 / 0.840 | 0.882 / **0.906** | 0.861 / 0.860 | 0.895 / 0.887 | 0.938 / 0.956 | **0.916** / 0.908 |
| GPT-4o | zero-shot | 0.906 / 0.902 | 0.826 / 0.876 | 0.864 / 0.878 | 0.952 / 0.945 | 0.868 / 0.917 | 0.908 / 0.919 |
|  | few-shot | **0.909 / 0.909** | 0.828 / 0.875 | **0.867 / 0.881** | **0.955 / 0.952** | 0.870 / 0.915 | 0.910 / **0.922** |
| GPT-5.2 | zero-shot | 0.762 / 0.758 | 0.855 / 0.874 | 0.806 / 0.796 | 0.835 / 0.824 | 0.938 / 0.954 | 0.884 / 0.867 |
|  | few-shot | 0.778 / 0.780 | 0.869 / 0.885 | 0.821 / 0.814 | 0.847 / 0.843 | 0.947 / 0.958 | 0.894 / 0.880 |
| GPT-5.4 | zero-shot | 0.731 / 0.736 | 0.882 / 0.902 | 0.799 / 0.792 | 0.786 / 0.785 | 0.949 / **0.960** | 0.860 / 0.844 |
|  | few-shot | 0.771 / 0.773 | **0.893 / 0.906** | 0.828 / 0.818 | 0.821 / 0.818 | **0.951** / 0.957 | 0.881 / 0.865 |
| LLaMA-3.3-70B | zero-shot | 0.836 / 0.844 | 0.823 / 0.877 | 0.829 / 0.847 | 0.888 / 0.888 | 0.873 / 0.924 | 0.880 / 0.892 |
|  | few-shot | 0.879 / 0.884 | 0.820 / 0.869 | 0.848 / 0.866 | 0.930 / 0.928 | 0.868 / 0.913 | 0.898 / 0.909 |
| MedGemma-text | zero-shot | 0.790 / 0.722 | 0.767 / 0.824 | 0.779 / 0.754 | 0.829 / 0.753 | 0.805 / 0.862 | 0.817 / 0.787 |
|  | few-shot | 0.845 / 0.781 | 0.773 / 0.827 | 0.807 / 0.791 | 0.887 / 0.815 | 0.812 / 0.864 | 0.848 / 0.825 |

Under the fuzzy span matching criterion, performance increased across all models. Few-shot GPT-4.1 reached the highest micro-F1, while few-shot GPT-4o maintained the highest macro-F1. Open-weight models also exhibited substantial performance improvement under this relaxed matching setting.

### Patient-Level Temporal Shift Distributions

As summarized in Table 2, each model generated 30 patient-level temporal shift configurations. Mean day-level and minute-level shifts showed high variability across models, with both positive and negative offsets observed. Specifically, GPT-4.1, GPT-4o, and LLaMA-3.3-70B demonstrated a general preference for negative day-level shifts, whereas MedGemma-text and GPT-5.4 averaged positive shifts. Results from the sample set largely mirrored those of the full dataset. While the overall direction of the shifts remained consistent, their magnitudes fluctuated. For instance, under the subset condition, GPT-5.2 exhibited a larger positive day-level shift, whereas GPT-5.4 produced a smaller one. Minute-level shifts followed a similar pattern of magnitude adjustment (e.g., larger positive offsets for GPT-5.4 and GPT-4.1, and more negative offsets for GPT-4o). The distribution of selected shifts across patients is expected and privacy-preserving, as it can reduce the risk of linkage through a shared temporal transformation. The large standard deviations therefore reflect intended shift value differences between patients rather than model inconsistency. In addition, differences between the full and sampled analyses are also expected because shift selection was conditioned on the prompt with temporal context provided to the model (Figure 1. Step 2).

**Table 2.** Distribution of Patient-Level Temporal Shift Values Selected by LLMs

| Models |  | Mean |  | Median |  | Std |  |
| --- | --- | --- | --- | --- | --- | --- | --- |
|  |  | Full | Sampled | Full | Sampled | Full | Sampled |
| GPT-5.4 | Day | 37.83 | 14.13 | 162.50 | 123.50 | 231.55 | 214.63 |
|  | Minute | 89.43 | 144.93 | 155.50 | 172.50 | 209.12 | 199.18 |
| GTP-5.2 | Day | 2.53 | 59.80 | -36.50 | 169.00 | 224.13 | 208.95 |
|  | Minute | 74.33 | 43.40 | 237.50 | 237.50 | 381.46 | 386.64 |
| GTP-4.1 | Day | -90.37 | -93.33 | -209.00 | -202.50 | 235.81 | 239.06 |
|  | Minute | 36.30 | 108.87 | 279.50 | 309.00 | 477.71 | 477.26 |
| GPT-4o | Day | -182.07 | -196.03 | -214.00 | -214.00 | 166.24 | 126.94 |
|  | Minute | -80.10 | -165.83 | -90.50 | -276.50 | 505.89 | 438.26 |
| Llama 3.3-70B | Day | -61.93 | -54.10 | -109.50 | -109.50 | 166.03 | 167.57 |
|  | Minute | 29.43 | 11.37 | 66.00 | 32.00 | 223.44 | 198.31 |
| MedGemma-text | Day | 50.30 | 58.70 | 155.00 | 169.00 | 221.57 | 255.03 |
|  | Minute | 178.33 | 176.43 | 155.50 | 254.00 | 320.15 | 365.15 |

**Figure 1.**
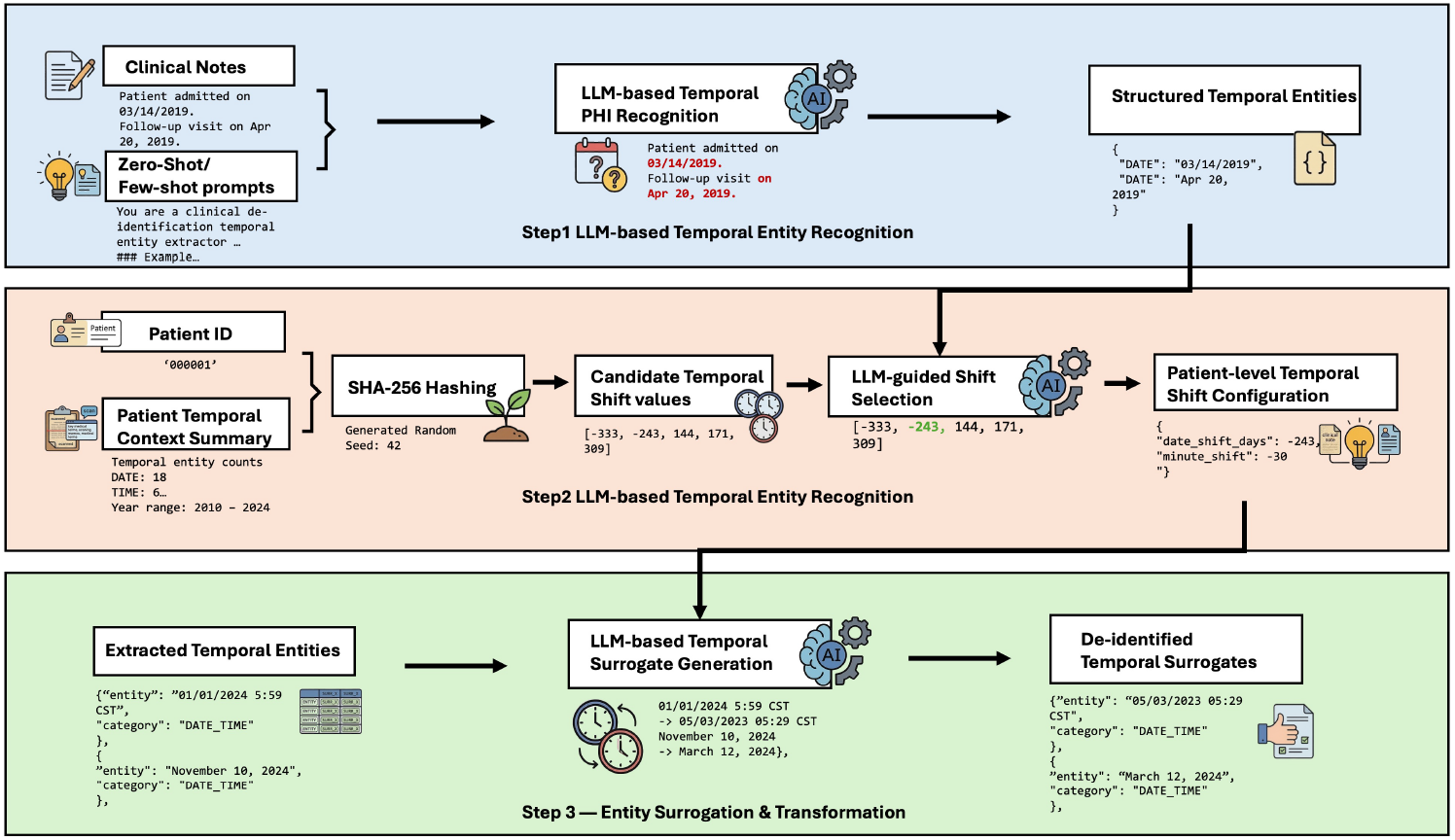
Overview of Temporal PHI De-identification Framework.

### Surrogation Generation Evaluation

Surrogate generation performance was evaluated based on format preservation, ordering integrity, and shift correctness (Table 3). Results from sample set was similar to the full dataset, suggesting that surrogate-generation performance was largely independent of differences in upstream extraction coverage.

**Table 3.**
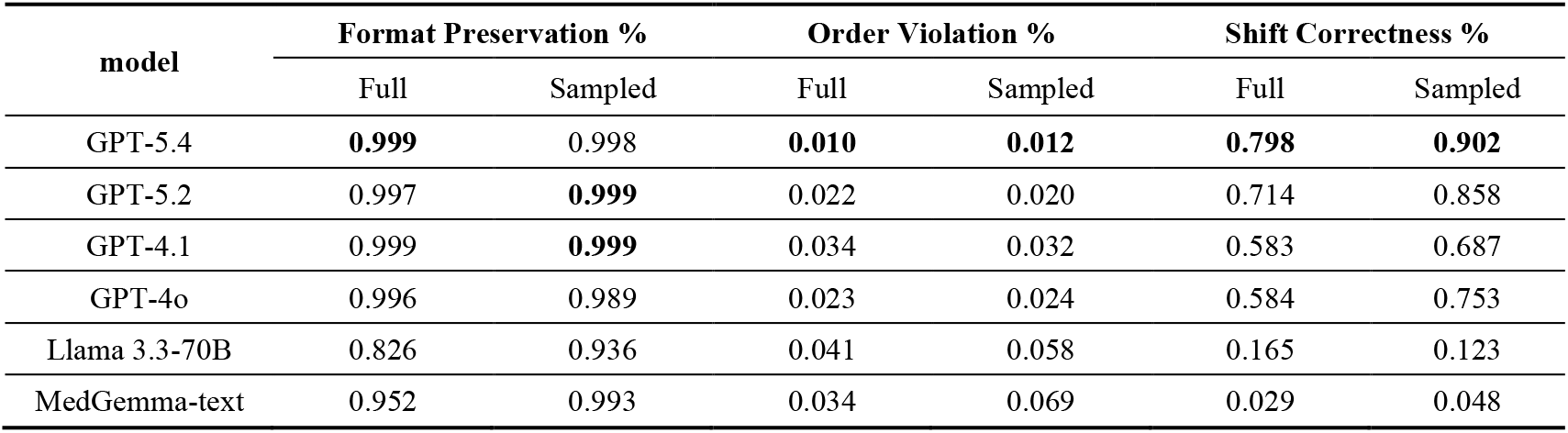
Performance of LLMs in Temporal Surrogate Generation

#### Formatting Consistency

Across both the full dataset and the sample set, format preservation was very high, exceeding 99% for most proprietary models. Open-weight models performed slightly lower, particularly LLaMA-3.3-70B (82.6% on the full set). Despite the high overall success rate, qualitative analysis revealed recurring formatting issues across models. Common issues included unprompted conversions from 24-hour to 12-hour formats (e.g., appending AM/PM markers to convert “MM/DD/YYYY HH:MM:SS” into “MM/DD/YYYY HH:MM:SS AM/PM”), introducing extra characters (e.g., leading newlines), and the irregularly normalization of informal expressions (e.g., expanding the abbreviation “5a” to “04:30 am”).

#### Ordering Integrity

GPT-5.4 achieved the lowest pairwise temporal order violation ratio (1.01% on the full set), followed closely by GPT-5.2 and GPT-4o. Error analysis revealed two recurring failure patterns: First, violations frequently involved partial date formats lacking year or day components (e.g., “M/D” or “M/YYYY”), which contributed the largest number of errors across leading models in the full entity set, including 413 errors in GPT-4.1, 290 in GPT-4o, 265 in GPT-5.2, and 154 in GPT-5.4. Second, order reversals also occurred when adjacent entities crossed month or year boundaries during shifting. This boundary-crossing pattern was observed in both the full and sampled sets.

#### Temporal Shifting Accuracy

Shift correctness showed lowest overall performance among the three metrics. When evaluated on the shared-entity subset, shift accuracy improved across all proprietary models. GPT-5.4 achieved the highest overall accuracy, increasing from 79.8% on the full set to 90.2% on the subset, followed by a similar improvement in GPT-5.2 (from 71.4% to 85.8%). In contrast, open-weight baselines struggled significantly across both settings (e.g., MedGemma-text achieved less than 5% correctness). In addition, analysis of residual temporal differences (Figure 2) demonstrated that shifting errors were not randomly distributed; rather, they consistently concentrated around the same offsets: ±1, ±30/31, and ±365 days. These recurring day-, month-, and year-scale deviations indicate a systemic limitation in the LLM models’ underlying calendar arithmetic.

**Figure 2.**
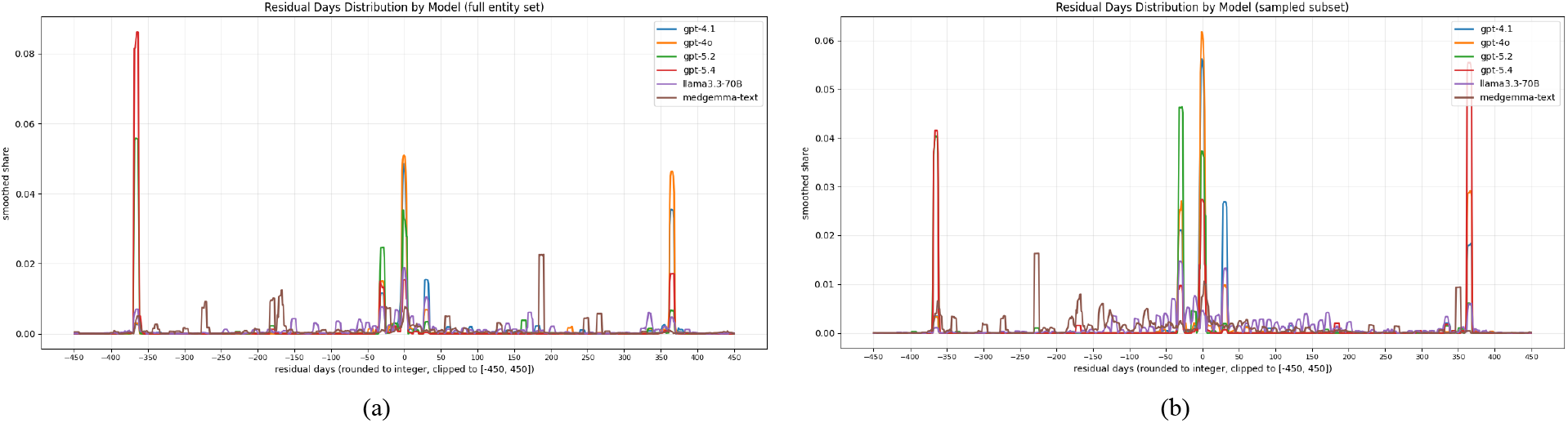
Residual Day Shift Distributions Across Models Under Full and Sampled Entity Sets.

## Discussion

Keeping the original temporal event order in clinical notes is important for causal analysis, modeling disease progression and longitudinal patient trajectories. Therefore, temporal entities must be accurately identified and shifted during de-identification to support multi-site data sharing. In this study, we systematically evaluated every component of an LLM-empowered framework for temporal PHI de-identification in real-world clinical notes. Our results demonstrate that LLMs can effectively extract temporal entities in clinical narratives across multiple models. However, model performance remained limited during temporal shifting. LLMs struggled with surrogate generation, particularly with applying the exact shifting value and preserving the temporal order.

### Strengths of LLM-based temporal PHI extraction

In the temporal PHI extraction evaluation, proprietary models showed consistently stronger performance. GPT-4o achieved the highest overall F1 score, while GPT-4.1 showed the highest recall among all models. Consistent with prior literature ^28-30^, the marginal performance gains observed from few-shot prompting suggest that modern LLMs already achieve robust inherent capabilities for zero-shot temporal entity recognition in clinical narratives. When we applied fuzzy matching, performance improved across all models, indicating that many errors stemmed from minor boundary variations rather than from complete entity-detection failures, which aligns with previous studies’ findings ^31,32^.

Despite these strong overall results, error analysis revealed consistent cross-model vulnerability. Irregular temporal expressions and time-only entities produced the highest false-negative and false-positive rates across most models. Such expressions often lack a clear structure and require contextual interpretation, posing a significant challenge for accurate LLM extraction. In contrast, numeric and text-based date formats were detected more consistently. These observations align with prior clinical NLP research showing that informal temporal expressions and relative time references remain challenging for automated extraction systems even when hybrid LLM-based approaches are used ^21,33^.

### Challenges in temporal surrogate generation

Generating temporal surrogates is distinct from entity detection; it requires accurate calendar arithmetic and the strict preservation of clinically meaningful intervals. While most LLMs successfully maintained the original formatting of temporal expressions (exceeding 99% accuracy), achieving chronological consistency was far more difficult. In our experiments, reasoning models demonstrated a clear advantage. Although GPT-5.4 and GPT-5.2 did not have the highest extraction scores, they consistently achieved the lowest temporal order violation ratios and the highest shift correctness. This performance divergence aligns with prior benchmark-based observations, indicating that pattern recognition and structured normalization or arithmetic tasks have different demands on language models ^30,34^.

Further analysis on shift errors showed that incorrect time shifts were not randomly distributed but clustered into common temporal patterns. The most frequent residuals were ±1 day, ±30/31 days, and ±365 days, corresponding to day-, month-, and year-scale deviations. These errors may reflect known limitations in temporal reasoning and date arithmetic in LLMs. Prior studies have shown that even state-of-the-art LLMs still struggle with temporal reasoning tasks, particularly when handling boundaries ^35^. One possible explanation for the observed residual patterns relates to how LLM tokenizers represent temporal expressions. Date strings are often fragmented into sub-tokens, disrupting the natural year-month-day structure. As noted by Bhatia et al. ^36^, LLMs must reconstruct this temporal information by stitching together these fragmented components, and high token fragmentation strongly correlates with reduced arithmetic accuracy. Additionally, temporal de-identification in clinical narratives involves handling incomplete or relative temporal expressions, which further complicates date shifting. The observed temporal-order violations and shift-value inaccuracies demonstrate that preserving the clinical timeline for reliable de-identification requires rigorous arithmetic handling that is beyond the basic string transformation capabilities of current LLMs.

### Future direction

Our multi-step evaluation results suggest that different LLMs present complementary strengths, indicating that temporal PHI de-identification tasks may benefit from a collaborative framework that leverages these strengths. One potential solution is to integrate a multi-agent pipeline into our framework, in which specialized models handle different components of the workflow ^37,38^. For instance, one model could focus on high-recall temporal entity extraction, while another reasoning or verification component performs temporal transformation and consistency checks. Such designs could enable systems to combine strong entity recognition capabilities with structured reasoning, thereby improving overall reliability for temporal PHI de-identification tasks. Furthermore, the predictable nature of the observed shifting errors presents a clear opportunity for targeted human-in-the-loop (HITL) integration. Instead of exhaustive manual review, future systems could automatically flag specific high-risk cases for human verification. This collaborative framework may provide a practical pathway for deploying LLM-based de-identification systems while maintaining the reliability required for clinical data sharing. Although this study focused on temporal PHI, future work should extend the framework to additional PHI categories such as names, locations, and organizations to further evaluate the applicability of LLM-based clinical de-identification. After applying the proposed de-identification framework to the sarcoma clinical notes dataset, we plan to release the de-identified dataset to support community benchmarking and future clinical research.

### Limitations

This study has several limitations. First, while our dataset contained over 28,000 temporal entities, the cohort was restricted to 30 sarcoma patients. This relatively small patient sample may limit the generalizability of our patient-level findings to broader clinical populations. Second, our dataset was collected from a limited number of healthcare institutions, which may not capture the full diversity of documentation styles, clinical workflows, and note structures across different healthcare systems. Third, due to privacy and institutional policy constraints, we were only able to evaluate proprietary models available within the institution’s approved cloud environment. In addition, the open-weight models were evaluated without task-specific fine-tuning; therefore, their performance may not reflect the potential of models trained on institution-specific clinical data. As a result, the comparison does not include all possible LLM variants or deployment settings. Finally, we did not conduct formal re-identification assessments or evaluate the impact of temporal de-identification on downstream tasks. Future work will address these limitations by evaluating larger and more diverse multi-institutional cohorts, assessing domain-adapted models, and examining privacy-utility trade-offs through re-identification and downstream-task evaluations.

### Conclusion

This study proposed and evaluated an LLM-based framework that integrates temporal entity extraction, patient-level temporal shift configuration, and surrogate generation to support temporal de-identification of real-world clinical notes. Using real-world clinical notes, we systematically evaluated multiple LLMs under zero-shot and few-shot prompting settings. Our findings demonstrate that modern LLMs are highly effective at extracting diverse temporal entities. However, generating reliable temporal surrogates remains a significant bottleneck. Error analysis revealed that model failures in shift-value correctness and temporal consistency are not random, but stem from systematic vulnerabilities in underlying calendar arithmetic and boundary handling. Ultimately, while LLMs show immense potential for automating PHI detection, preserving the chronological integrity of clinical timelines requires moving beyond standalone generative models toward specialized, collaborative multi-agent systems and targeted human-in-the-loop workflows.

## Data Availability

All data produced in the present study are available upon reasonable request to the authors.

## Acknowledgments

Research reported in this publication was supported by the National Library of Medicine of the National Institutes of Health under award numbers R01LM011934, the National Human Genome Research Institute under award number R01HG012748, the National Institute of Aging under award number R01AG072799, the Cancer Prevention Institute of Texas (CPRIT) under award number RR230020, and the Food and Drug Administration (FDA) under award number U01FD008720. The content is solely the responsibility of the authors and does not necessarily represent the official views of the National Library of Medicine, the National Human Genome Research Institute, the National Institutes of Health, or the State of Texas. We thank Wanjing Wang and Huipeng Liu for assistance with data preparation and gold-standard annotation, and Qiuhao Lu for support with data cleaning.

